# MDA5 variants trade antiviral activity for protection from autoimmune disease

**DOI:** 10.1101/2024.10.04.24314884

**Authors:** Chris Wallace, Rahul Singh, Yorgo Modis

## Abstract

Loss-of-function variants in MDA5, a key sensor of double-stranded RNA from viruses and retroelements, have been associated with protection from type 1 diabetes (T1D) in genome-wide association studies (GWAS). MDA5 loss-of-function variants have also been reported to increase the risk of inflammatory bowel disease (IBD). Whether these associations are linked or extend to other diseases remains unclear. Here, fine-mapping analysis of four large GWAS datasets shows that T1D-protective loss-of-function MDA5 variants also protect against psoriasis and hypothyroidism, while increasing the risk of IBD. The degree of autoimmune protection and IBD risk were linearly proportional. The magnitudes of the odds ratios for autoimmune protection and IBD risk were larger for rare MDA5 variants than for common variants, which were differentially expressed in different geographic populations. Our analysis suggests MDA5 genetic variants offer a direct fitness trade-off between viral clearance and autoimmune tissue damage.

## Introduction

Innate immune responses must be sensitive enough to detect infection yet specific enough to avoid activation by cellular components. A key innate immune receptor for cytosolic double-stranded RNA (dsRNA), a potent signature of viral infection, is MDA5 (Berke & Modis, 2012; Kato *et al*, 2006; Peisley *et al*, 2011; Yu *et al*, 2021; Yu *et al*, 2018). Recognition of dsRNA by MDA5 induces a potent IFN-β response (Berke & Modis, 2012; Peisley *et al*., 2011). The ATPase activity and dsRNA binding cooperativity of MDA5 confer the necessary sensitivity and specificity of MDA5-dsRNA recognition (Singh *et al*, 2024b; Yu *et al*., 2021; Yu *et al*., 2018). Mutations in the gene encoding MDA5, *IFIH1*, can perturb the balance between sensitive and specific dsRNA recognition. *IFIH1* is a hotspot for natural variants with clinical associations. Approximately 40 gain-of-function missense variants have been associated with interferonopathies (Rice *et al*, 2020; Rodero & Crow, 2016; Rutsch *et al*, 2015). These variants promote formation of MDA5 signaling complexes, including on endogenous dsRNAs, by either increasing the RNA binding affinity of MDA5 or inhibiting its ATP-dependent proofreading activity (Garau *et al*, 2019; Rice *et al*., 2020; Yu *et al*., 2021). Loss-of-function MDA5 variants cause recurrent infections (Lamborn *et al*, 2017), and have been reported to contribute to inflammatory bowel disease (IBD), including ulcerative colitis and Crohn’s disease (Adiliaghdam *et al*, 2022; Cananzi *et al*, 2021). GWAS data have also associated loss-of-function MDA5 variants with reduced risk of developing certain autoimmune diseases, most notably T1D (Downes *et al*, 2010; Gorman *et al*, 2017; Liu *et al*, 2009; Nejentsev *et al*, 2009; Smyth *et al*, 2006; Vasseur *et al*, 2011). Specifically, variants E627* (rs35744605), R843H (rs3747517), I923V (rs35667974), and T946A (rs1990760) have been identified as T1D-protective (de Azevedo Silva *et al*, 2015; Gorman *et al*., 2017; Jermendy *et al*, 2018; Liu *et al*., 2009; Nejentsev *et al*., 2009; Smyth *et al*., 2006; Vasseur *et al*., 2011). The E627* and I923V variants are rare, with allele frequencies of 1-2%, while R843H and T946A are common. These common variants are differentially expressed in different geographic populations. The T946A variant is present in 70-80% of Africans and Asians but only 30-50% of Caucasians (de Azevedo Silva *et al*., 2015; Gorman *et al*., 2017; Liu *et al*., 2009; Nejentsev *et al*., 2009; Vasseur *et al*., 2011). Similarly, the R843H variant is found in 70% of Asians but only 30-40% of Caucasians and Africans (de Azevedo Silva *et al*., 2015; Gorman *et al*., 2017; Liu *et al*., 2009; Nejentsev *et al*., 2009; Vasseur *et al*., 2011). Most human *IFIH1* reference sequences contain the alleles that are most common in Asians (Ala946/His843). These coding variants inhibit the formation of MDA5-dsRNA signaling complexes (Gorman *et al*., 2017; Shigemoto *et al*, 2009) by reducing the RNA binding affinity of MDA5 or hyperactivating its ATPase activity (Singh *et al*, 2024a). Other loss-of-function variants are splice donor variants that reduce splicing efficiency and hence mRNA levels (Downes *et al*., 2010).

A robust clinical link has emerged between T1D onset and recent infection with RNA viruses, in particular coxsackieviruses and other enteroviruses (Yeung *et al*, 2011). T1D patients have more frequent and persistent enterovirus infections, which precede the appearance of prediabetic markers, including autoantibodies (Hyoty & Taylor, 2002). MDA5 recognizes RNA from *Picornaviridae*, including enteroviruses (Kato *et al*., 2006; Wang *et al*, 2010), which have evolved mechanisms to suppress IFN-β transcription (Hato *et al*, 2007; Visser *et al*, 2019). MDA5-induced inflammation in the pancreas following rotavirus infection contributes to autoimmune destruction of pancreatic β-cells (Dou *et al*, 2017). Therefore, a plausible hypothesis is that MDA5-dependent inflammation following viral infection can trigger autoimmune β-cell killing.

Gastrointestinal viral infection has also been linked to IBD onset(Axelrad *et al*, 2019). Intestinal cells from IBD patients harboring loss-of-function MDA5 variants had elevated viral loads and were compromised in their ability to maintain epithelial barrier integrity upon further exposure to the enteric virome (Adiliaghdam *et al*., 2022). Hence, the frequency and clinical phenotypes of MDA5 loss-of-function variants in IBD patients suggest that MDA5 deficiency contributes to the induction of IBD (Adiliaghdam *et al*., 2022; Cananzi *et al*., 2021), due in part to increased exposure and susceptibility to viral infection (Adiliaghdam *et al*., 2022; Lamborn *et al*., 2017).

## Results and discussion

Loss-of-function MDA5 variants have been associated with recurrent infection (Lamborn *et al*., 2017), T1D protection (Downes *et al*., 2010; Gorman *et al*., 2017; Liu *et al*., 2009; Nejentsev *et al*., 2009; Smyth *et al*., 2006; Vasseur *et al*., 2011), and IBD risk (Adiliaghdam *et al*., 2022; Cananzi *et al*., 2021). However, whether these associations are linked, or whether they apply to autoimmune disease more broadly, has not been explored. We addressed these questions by utilizing larger, more recent GWAS studies to conduct a broader, more sensitive survey of immune-mediated disease associations involving MDA5 variants. For our analysis, we selected GWAS datasets to maximize the likelihood of accurate fine mapping – balancing the need for large sample size, minimizing the use of imputed genotypes, and using ancestries that best matched the reference panel used to derive linkage disequilibrium (LD) estimates (Karczewski *et al*, 2024; Liu *et al*, 2015; Robertson *et al*, 2021; Tsoi *et al*, 2017) (see **Materials and methods**). By reviewing associations with a reported lead T1D-associated variant, I923V (rs35667974) (Nejentsev *et al*., 2009) we identified psoriasis, hypothyroidism, Crohn’s disease, and ulcerative colitis as additional associated diseases. Fine-mapping analysis using GWAS summary data for these five diseases identified four variants likely to be causally associated with a subset of the diseases, with considerable overlap of variants between diseases (**Fig. 1, Table I**). Two of these variants were coding variants, I923V and T946A, and two were splice donor variants, rs35337543 and rs35732034. All four variants were previously associated with T1D protection (Downes *et al*., 2010; Gorman *et al*., 2017; Liu *et al*., 2009; Nejentsev *et al*., 2009; Smyth *et al*., 2006; Vasseur *et al*., 2011).

**Fig 1.**
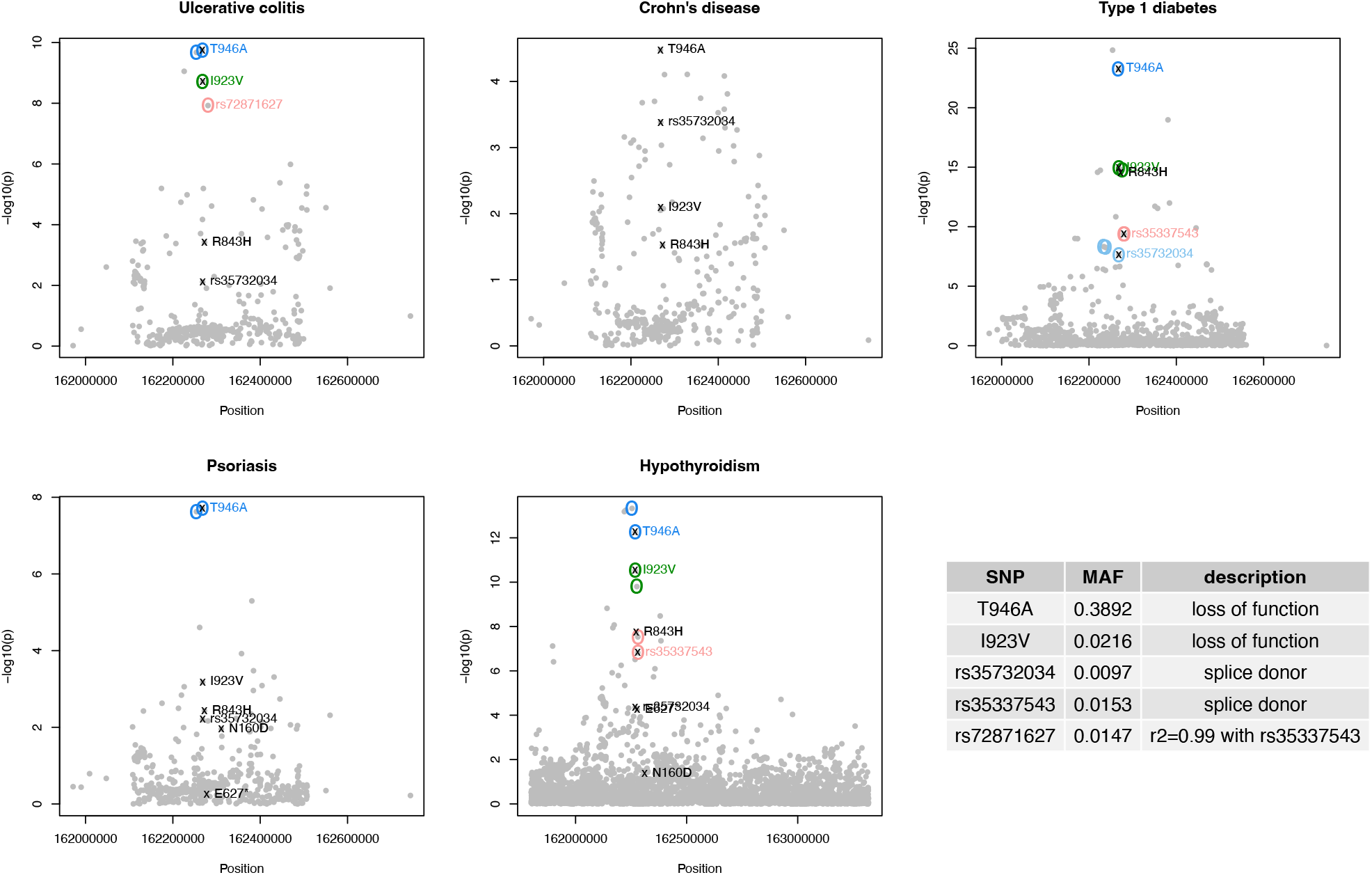
Manhattan plots of fine-mapped variants for five diseases. Colored circles indicate the fine mapping sets identified, and colored text labels the most likely causal SNP in each set, selected due its known function on MDA5. Known loss-of-function SNPs that were not in any fine mapping set are marked with “x” and a black label. Lower right, minor allele frequency (MAF) and function of fine-mapped SNPs.

**Table 1.** Marginal association and fine-mapping analysis results.

| Variant | SNP | Location | Allele | EAF | Disease | lnOR ± std. err. | OR | P | P<sub>Bonferroni</sub> | Causal<sup>‡</sup> |
| --- | --- | --- | --- | --- | --- | --- | --- | --- | --- | --- |
| I923V | rs35667974 | 2:162268127 | T>C | 0.022 | T1D | -0.493 ± 0.062 | 0.611 | 1.21×10<sup>-15</sup> | 2.91×10<sup>-14</sup> | T |
| | | | | | Psoriasis | -0.440 ± 0.129 | 0.644 | 6.68×10<sup>-4</sup> | 0.0160 | F |
| | | | | | Hypothy. | -0.254 ± 0.038 | 0.776 | 2.98×10<sup>-11</sup> | 7.16×10<sup>-10</sup> | T |
| | | | | | Crohn's | 0.121 ± 0.046 | 1.129 | 8.08×10<sup>-3</sup> | 0.1938 | F |
| | | | | | UC | 0.276 ± 0.046 | 1.318 | 1.94×10<sup>-9</sup> | 4.64×10<sup>-8</sup> | T |
| Splice1 | rs35732034 | 2:162268086 | C>T | 0.010 | Psoriasis | -0.693 ± 0.253 | 0.500 | 6.25×10<sup>-3</sup> | 0.1500 | F |
| | | | | | T1D | -0.464 ± 0.083 | 0.629 | 2.29×10<sup>-8</sup> | 5.50×10<sup>-7</sup> | T |
| | | | | | Hypothy. | -0.230 ± 0.056 | 0.794 | 4.47×10<sup>-5</sup> | 1.07×10<sup>-3</sup> | F |
| | | | | | UC | 0.180 ± 0.067 | 1.197 | 7.70×10<sup>-3</sup> | 0.1848 | F |
| | | | | | Crohn's | 0.226 ± 0.064 | 1.253 | 4.15×10<sup>-4</sup> | 9.96×10<sup>-3</sup> | F |
| Splice2* | rs72871627 | 2:162280432 | A>G | 0.015 | Psoriasis | -0.541 ± 0.200 | 0.582 | 6.81×10<sup>-3</sup> | 0.1634 | F |
| | | | | | T1D | -0.458 ± 0.073 | 0.633 | 4.19×10<sup>-10</sup> | 1.00×10<sup>-8</sup> | T |
| | | | | | Hypothy. | -0.253 ± 0.046 | 0.777 | 2.94×10<sup>-8</sup> | 7.05×10<sup>-7</sup> | T |
| | | | | | Crohn's | 0.072 ± 0.056 | 1.074 | 0.1990 | 1 | F |
| | | | | | UC | 0.308 ± 0.054 | 1.360 | 1.19×10<sup>-8</sup> | 2.85×10<sup>-7</sup> | T |
| Splice2 | rs35337543 | 2:162279995 | C>G | 0.015 | T1D | -0.458 ± 0.073 | 0.633 | 4.19×10<sup>-10</sup> | ND | T |
| | | | | | Hypothy. | -0.236 ± 0.045 | 0.790 | 1.51×10<sup>-7</sup> | ND | T |
| E627* | rs35744605 | 2:162277580 | C>A | 0.007 | Hypothy. | -0.273 ± 0.068 | 0.761 | 5.63×10<sup>-5</sup> | 1.35×10<sup>-3</sup> | F |
| | | | | | Psoriasis | -0.210 ± 0.372 | 0.810 | 0.5717 | 1 | F |
| T946A | rs1990760 | 2:162267541 | T>C | 0.389 | Psoriasis | -0.192 ± 0.034 | 0.825 | 1.95×10<sup>-8</sup> | 4.68×10<sup>-7</sup> | T |
| | | | | | T1D | -0.132 ± 0.013 | 0.876 | 5.57×10<sup>-24</sup> | 1.34×10<sup>-22</sup> | T |
| | | | | | Hypothy. | -0.080 ± 0.011 | 0.923 | 5.61×10<sup>-13</sup> | 1.35×10<sup>-11</sup> | T |
| | | | | | Crohn's | 0.053 ± 0.013 | 1.055 | 3.36×10<sup>-5</sup> | 8.07×10<sup>-4</sup> | F |
| | | | | | UC | 0.086 ± 0.013 | 1.089 | 1.78×10<sup>-10</sup> | 4.27×10<sup>-9</sup> | T |
| R843H | rs3747517 | 2:162272314 | C>T | 0.276 | T1D | -0.111 ± 0.014 | 0.895 | 2.94×10<sup>-15</sup> | ND | F |
| | | | | | Psoriasis | -0.103 ± 0.035 | 0.903 | 3.72×10<sup>-3</sup> | ND | F |
| | | | | | Hypothy. | -0.068 ± 0.012 | 0.934 | 1.85×10<sup>-8</sup> | ND | F |
| | | | | | Crohn's | 0.030 ± 0.014 | 1.031 | 0.0301 | ND | F |
| | | | | | UC | 0.051 ± 0.014 | 1.053 | 3.85×10<sup>-4</sup> | ND | F |
| N160D | rs74162075 | 2-162310909 | T>C | 0.001 | Psoriasis | -2.057 ± 0.811 | 0.128 | 0.0111 | 0.2674 | F |
| | | | | | Hypothy. | -0.270 ± 0.133 | 0.763 | 0.0427 | 1 | F |

EAF, effect allele frequency; OR, odds ratio.
\*rs72871627 was used as a tag of rs35337543 because the latter was not available in all GWAS datasets.
<sup>‡</sup>Causality was inferred from presence of the SNP in a credible set in fine mapping analysis.
Dashed lines indicate that the SNPs above and below the line are in LD with each other.

Given the overlaps between diseases, and the small *P* values observed for fine-mapped variants in one disease when considering another, we called associated variants by considering marginal association at any of these variants or other known loss-of-function variants not in LD with any of the index variants using a stringent Bonferroni correction. This identified associations of a fifth variant, E627* (rs35744605), with psoriasis, and hypothyroidism (**Fig. 1**). While T946A is common, the other variants are rare (allele frequency below 3%). Fine-mapping analysis did not find evidence for causality for variants E627* and R843H (**Table I**). However, for E627* the lack of evidence of causality is likely due to its low allele frequency (0.7%), which limits power in genetics studies. In contrast, association of R843H with T1D protection is likely attributable to LD with the co-occurring causal T946A allele (r^2^ = 0.60), as previously reported (Gorman *et al*., 2017; Liu *et al*., 2009; Nejentsev *et al*., 2009). Supporting these conclusions, our accompanying biochemical and structural study of T1D-protective MDA5 variants shows that the E627* variant lacks signaling activity due to a loss of RNA binding affinity, whereas the R843H substitution had no effect on the structural, biochemical or signaling activities of MDA5 (Singh *et al*., 2024a).

Comparing associations across diseases, we found that all five associated variants offered protection against T1D, psoriasis and hypothyroidism, but increased the risk of ulcerative colitis and Crohn’s disease (**Fig. 2a**). Remarkably, a strict correlation was observed between protection and risk (**Fig. 2b**). A statistical test of proportionality (see **Materials and methods**) concluded that changes in risk were linearly proportional across variants (**Fig. 2c**). Additionally, the magnitudes of the odds ratios were larger for the rare variants than for the common variants. Thus, the rarest variant, I923V, conferred the greatest degree of T1D protection and the greatest risk of IBD, whereas the most common variant, T946A, was associated with the least T1D protection and IBD risk (**Fig. 2a-b**). Notably, the T946A variant, while common in all populations, is the major allele in Asian and African populations but the minor allele in Caucasians.

**Figure 2.**
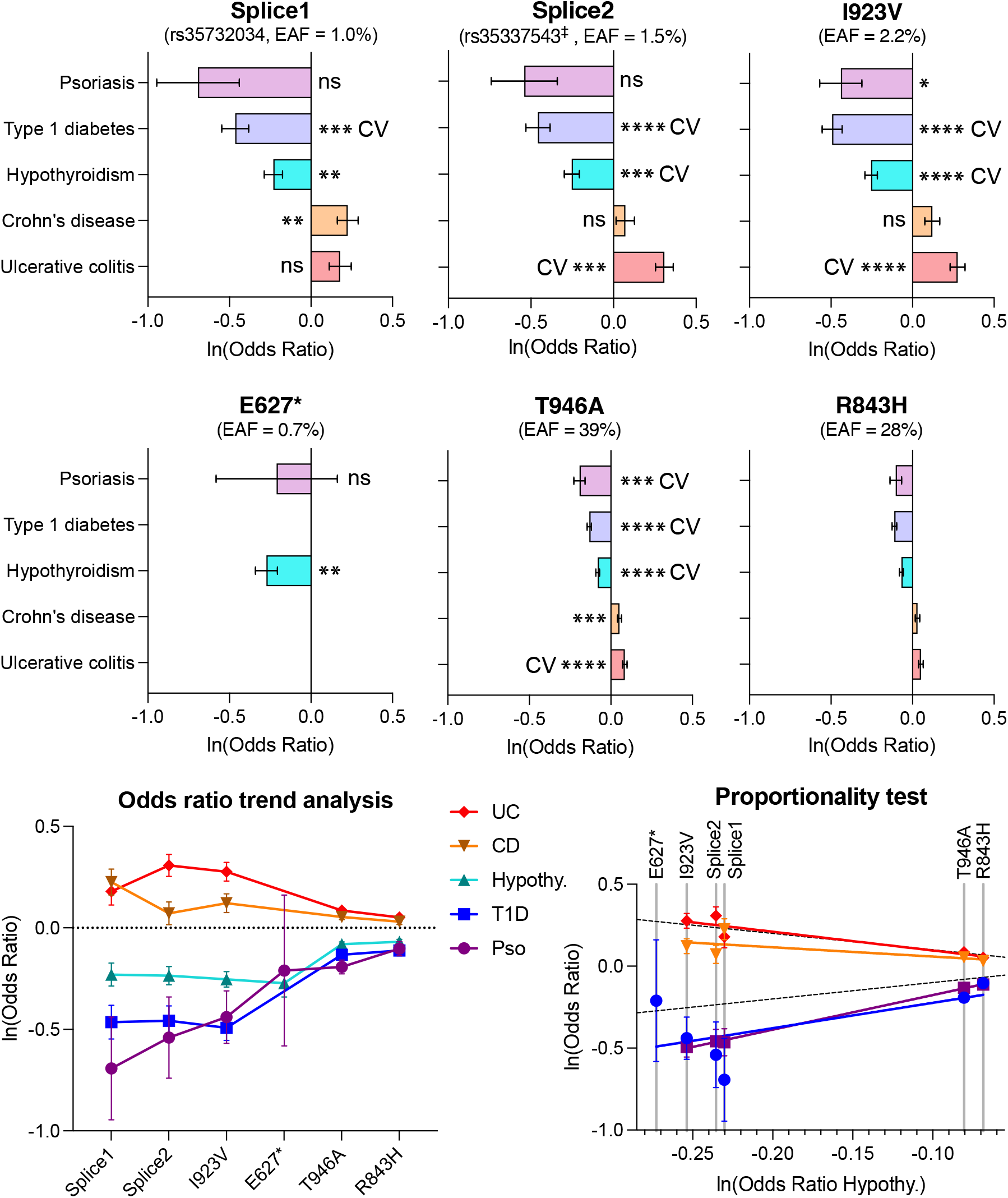
GWAS meta-analysis of loss-of-function MDA5 variants. **(a)** Effect sizes expressed in terms of the odd ratio for selected MDA5 loss-of-function variants. ^‡^Because rs35337543 was only available in two GWAS datasets, rs72871627 (r^2^ = 0.973) was used as a tag for rs35337543. EAF, effect allele frequency. Error bars, SEM. ns, P > 0.05; *, P < 0.05; **, P < 0.01; ***, P < 0.001; ****, P < 5**×**10^-8^ (genome-wide significance), where P is the Bonferroni-adjusted P value from marginal association analysis. CV, likely causal variant present in a credible set in fine-mapping analysis. R843H was included in fine-mapping analysis but excluded from marginal association analysis. See **Table I**. **(b)** Odds ratio plot for variants and diseases in *(a)*. **(c)** Estimated effect sizes in hypothyroidism compared to the other diseases. Hypothyroidism was selected because it was the largest dataset for which all SNPs were available. Dashed lines, identity and negative identity lines.

Our GWAS meta-analysis uncovers a direct correlation between protection from autoimmune-related diseases (T1D, psoriasis and hypothyroidism) and increased risk of IBD. The implication is that calibration of MDA5-dependent antiviral signaling offers a fundamental fitness trade-off. Loss-of-function MDA5 variants protect from autoimmune tissue damage, including to pancreatic β-cells leading to T1D, but increase the risk of inflammatory tissue damage from persistent infection, including by enteric viruses in intestinal epithelia leading to IBD. This model predicts that gain-of-function MDA5 variants protect from chronic inflammation and IBD by ensuring infections are cleared but do so at the cost of increasing the risk of T1D and other autoimmune diseases.

## Materials and methods

### GWAS meta-analysis

GWAS summary data for a reported lead T1D-associated variant, I923V (rs35667974) were downloaded for the following diseases: T1D (16,000 T1D cases, 25,000 controls) (Robertson *et al*., 2021); IBD (5,956 Crohn’s disease cases, 6,968 ulcerative colitis cases, 21,770 controls) (Liu *et al*., 2015); psoriasis (19,032 cases, 286,769 controls) (Tsoi *et al*., 2017); and hypothyroidism (20,563 cases, 399,910 controls) (Karczewski *et al*., 2024). Data were downloaded from the GWAS catalog from the following repositories: ulcerative colitis, https://ftp.ebi.ac.uk/pub/databases/gwas/summary_statistics/GCST003001-GCST004000/GCST003045/harmonised/26192919-GCST003045-EFO_0000729.h.tsv.gz (ImmunoChip); Crohn’s disease, https://ftp.ebi.ac.uk/pub/databases/gwas/summary_statistics/GCST003001-GCST004000/GCST003044/harmonised/26192919-GCST003044-EFO_0000384.h.tsv.gz (ImmunoChip); T1D, https://ftp.ebi.ac.uk/pub/databases/gwas/summary_statistics/GCST90013001-GCST90014000/GCST90013445/GCST90013445_buildGRCh38.tsv (ImmunoChip); Psoriasis: http://ftp.ebi.ac.uk/pub/databases/gwas/summary_statistics/GCST005001-GCST006000/GCST005527/harmonised/23143594-GCST005527-EFO_0000676.h.tsv.gz (Meta analysis); Hypothyroidism: https://pan-ukb-us-east-1.s3.amazonaws.com/sumstats_flat_files/categorical-20002-both_sexes-1453.tsv.bgz (genome wide SNP chip). These studies were selected to balance the need for large studies and those most likely to give accurate results in fine mapping – i.e. those which minimized imputation and most closely matched the ancestry of our reference LD panel.

Alleles were aligned to the UK Biobank as a common reference. Reference LD matrices were estimated from 40,000 European subjects from UK Biobank. Fine mapping was performed with a variant of the Sum of Single Effects (SuSiE) (Wang *et al*, 2020) model, as implemented in susieR (Zou *et al*, 2022).

rs72871627 was selected as a tag (r^2^ = 0.99) of the splice donor variant rs35337543 because rs72871627 was available in all GWAS datasets whereas rs35337543 was only available in the T1D GWAS dataset.

Tests of proportional effects between pairs of diseases were performed across all variants in Table I using colocPropTest [https://cran.r-project.org/package=colocPropTest] which implements the test of proportionality as previously described (Wallace, 2013).

## Data Availability

This study is a meta-analysis of previously published data. All code to reproduce this analysis is available at Zenodo.org [https://doi.org/10.5281/zenodo.12771481].

## Competing Interests

Y.M. is a consultant for Related Sciences LLC. C.E.W. is a part time employee at GSK. GSK was not involved in this study. R.S. has no competing interests or conflicts to declare.

## Author Contributions

Conceptualization, C.E.W., R.S. and Y.M.; Methodology, C.E.W.; Formal Analysis, C.E.W. and Y.M.; Funding Acquisition, Y.M.; Project Administration, Y.M.; Software, C.E.W.; Supervision, Y.M.; Writing – Original Draft, Y.M.; Writing – Review & Editing, C.E.W., R.S. and Y.M.

## Acknowledgements

We thank Louise Modis, Tetsuo Hasegawa, and the Modis group for useful discussions. This work was supported by the Wellcome Trust [217191/Z/19/Z to Y.M.] and [WT220788 to C.E.W], and the Medical Research Council [MC_UU_00040/01 to C.E.W.].

## Notes

### Author Declarations

Source data were openly available before the initiation of the study. Data can be downloaded from the GWAS catalog from the EBI repositories using the links provided in the Data Availability Links section.

### Summary of Updates

Additional funding sources and competing interests were added to the Funding statement and Competing interests statement, respectively. Minor edits to the main text to correct typos or clarify the text.

